# Glutamate levels across deep brain structures in patients with a psychotic disorder and its relation with cognitive functioning

**DOI:** 10.1101/2021.02.01.21250977

**Authors:** Tommy A.A. Broeders, Alex A. Bhogal, Lisan M. Morsinkhof, Menno M. Schoonheim, Christian H. Röder, Mirte Edens, Dennis W.J. Klomp, Jannie P. Wijnen, Christiaan H. Vinkers

**Affiliations:** Department of Radiology, University Medical Center Utrecht, Utrecht, The Netherlands; Department of Anatomy & Neurosciences, Amsterdam UMC (location VU University Medical Center), Amsterdam, The Netherlands; Department of Psychiatry, Amsterdam UMC (location VU University Medical Center)/GGZ inGeest, Amsterdam, The Netherlands; Department of Psychiatry, Brain Center Rudolf Magnus, University Medical Center Utrecht, Utrecht, The Netherlands

**Keywords:** magnetic resonance spectroscopic imaging (MRSI), proton spectroscopy, glutamate, psychotic disorder, cognition, psychomotor speed

## Abstract

Patients with psychotic disorders often show prominent cognitive impairment. Glutamate seems to play a prominent role, but knowledge on its role in deep gray matter regions is limited and previous studies have yielded heterogeneous results. The aim was to evaluate glutamate levels within deep gray matter structures in patients with a psychotic disorder in relation to cognitive functioning, using advanced spectroscopic acquisition, reconstruction and post-processing techniques. A 7 tesla MRI scanner combined with a unique lipid suppression coil and subject specific water signal suppression pulses were used to acquire high-resolution magnetic resonance spectroscopic imaging data. Anatomical scans were used to perform tissue fraction correction and registration to a standard brain for group comparison in specifically delineated brain regions. The brief assessment of cognition in schizophrenia was used to evaluate cognitive status. Average glutamate levels across deep gray matter structures (i.e. caudate, pallidum, putamen, and thalamus) in patients with a psychotic disorder (n=16, 4 females) were lower compared to healthy controls (n=23, 7 females). Stratified analyses showed lower glutamate levels in the caudate and putamen but not in the pallidum or thalamus. Average glutamate levels across deep gray matter structures were positively correlated with cognition, particularly to psychomotor speed. We find reduced glutamate levels across deep brain structures such as the caudate and putamen in patients with a psychotic disorder that are linked to psychomotor speed. Our results underscore the potential role of detailed *in vivo* glutamate assessments to understand cognitive deficits in patients with psychotic disorders.

## INTRODUCTION

Patients with psychotic disorders often show prominent cognitive deficits that greatly impact their quality of life and these deficits determine long-term outcomes more than positive symptoms (i.e. hallucinations and delusions).^1, 2^ These cognitive deficits often precede psychosis onset and persist after the acute phase of the illness and despite pharmacological treatment.^2-4^ However, the underlying pathophysiology of cognitive deficits in psychotic disorders is currently poorly understood. Glutamatergic dysfunction is involved in the development and course of psychotic disorders, and it might be involved in the etiology of cognitive deficits as well.^5, 6^ Assessing glutamate levels may be important for understanding these cognitive deficits. ^7^ The main technique for directly assessing *in vivo* glutamate levels is proton magnetic resonance spectroscopy (^1^H-MRS).^8^ The majority of ^1^H-MRS studies have focused on frontal or medial cortical areas, and, to a lesser extent, the basal ganglia and the thalamus.^9, 10^ Accordingly, a recent review indicated a paucity in studies targeting subcortical regions where glutamatergic deficits may be pronounced.^11^ Glutamate levels in subcortical regions may be particularly important for understanding cognition in schizophrenia, but also disease progression in individuals at clinical high risk.^12, 13^

Over the past decades, an increasing number of studies have used ^1^H-MRS to quantify glutamate, glutamine, and their combined signal (Glx) in patients with psychotic disorders.^14, 15^ A recent meta-analysis, summarizing ^1^H-MRS studies at 1.5 to 4 tesla in schizophrenia, reported elevated Glx,^16^ while earlier meta-analyses reported more heterogeneous results.^9, 10^ This variability may at least partially arise from limited signal-to-noise ratios and effects relating to overlapping resonances in the glutamate levels at lower magnetic field strength, including the separation of glutamate and glutamine resonances. A major consideration in the interpretation of MRS studies is that spectra are typically obtained unilaterally in one large predetermined voxel of interest (often 8 cm^3^), often averaging gray (GM) and white matter (WM) signals. Because of this, the question remains *which* deep brain structure to target when investigating disease-mediated changes in glutamate levels and their relation with cognitive deficits. Ultra-high field approaches (7T and higher) may overcome such issues and have generally reported reduced or comparable glutamate levels in schizophrenia patients.^17-21^

The aim of this study was therefore to acquire MRSI data of deep brain glutamate levels in patients with a psychotic disorder and examine its relation to cognitive deficits. To allow the simultaneous targeting of multiple deep brain regions in a single acquisition, we used a high resolution 2D magnetic resonance spectroscopic imaging (MRSI) approach at 7 Tesla. Novel methodological combinations were applied to enable reliable group analyses on glutamate levels across accurately delineated deep brain regions. In line with previous studies at ultra-high field, we hypothesized that patients with a psychotic disorder would show reduced glutamate values which would be regionally specific and associated with cognitive dysfunction.

## Materials and Methods

### General

Subjects were included at the University Medical Center Utrecht (UMCU). Inclusion criteria were the presence of a psychotic disorder as assessed using the mini-international neuropsychiatric interview (MINI).^22^ Patients were excluded with a drug-induced psychosis or an affective disorder with psychotic features. Individuals without lifetime neurological or psychiatric conditions were recruited as healthy controls. Individuals without lifetime neurological or psychiatric conditions were recruited as healthy control. Patients were excluded with a drug-induced psychosis or an affective disorder with psychotic features as assessed with the MINI.^22^ All participants were requested to abstain from recreational drugs in the two week period prior to the study, and alcohol within 24 hours prior to the study. This study was approved by the medical research ethics committee of UMCU and written informed consent was obtained from all subjects. The experiments were performed according to the guidelines and regulations of the WMO (Wet Medisch Wetenschappelijk Onderzoek).

### Psychiatric evaluation and medication

The MINI was administered before study participation by TAAB, LM and ME, trained by an experienced psychiatrist. The participant was asked to list the names and doses of any antipsychotic medication they were taking.

### Cognitive assessment

The brief assessment of cognition in schizophrenia (BACS) was used for evaluation of cognitive status, and administered before MRI scanning on the same day.^23^ The BACS included eight items covering six domains; verbal memory, working memory, motor speed, verbal fluency (three items), attention and processing speed, and executive function. The raw scores of each item were transformed to z-scores based on the distribution of the healthy controls. The three items for verbal fluency were averaged, resulting in six z-scores per participant indicating the cognitive score per domain. Finally, all six z-scores were averaged to quantify average cognition.

### MRI data

MRSI methodology was used as previously published.^24^ In short, data were acquired using a 7 tesla MR scanner (Phillips, Best, NL) equipped with a dual-transmit head coil and 32-channel receive coil (Nova Medical, Wilmington Ma, USA). Second order image-based shimming was performed for all acquisitions. Suppression of extra-cranial lipid signals (and therefore significant lipid contamination artefacts)^25^ was achieved using an external lipid suppression coil.^26^ MRSI data was acquired using a slice-selective, free-induction decay sequence^27^ with the following parameters: TE/TR = 2.5/300 ms, FOV = 220×220 mm, acquisition matrix = 44×44, resolution 5×5×10 mm^3^, BW = 3000 Hz, samples = 512, signal averages = 2, elliptical k-space, scan duration = 10 m 59 s, subject specific spiral in-out spectral-spatial water suppression pulses,^28^ flip angle = 35°. Water unsuppressed MRSI data was acquired for zeroth order phase and eddy current correction with adapted parameters: acquisition matrix = 22×22, resolution 10×10×10mm^3^, signal averages = 1, scan duration = 1m 54s. Two adjacent 10mm MRSI slices (20mm slab) were acquired axially to intersect deep GM nuclei located adjacent to the ventricles (**Figure 1A**).

**Figure 1A:**
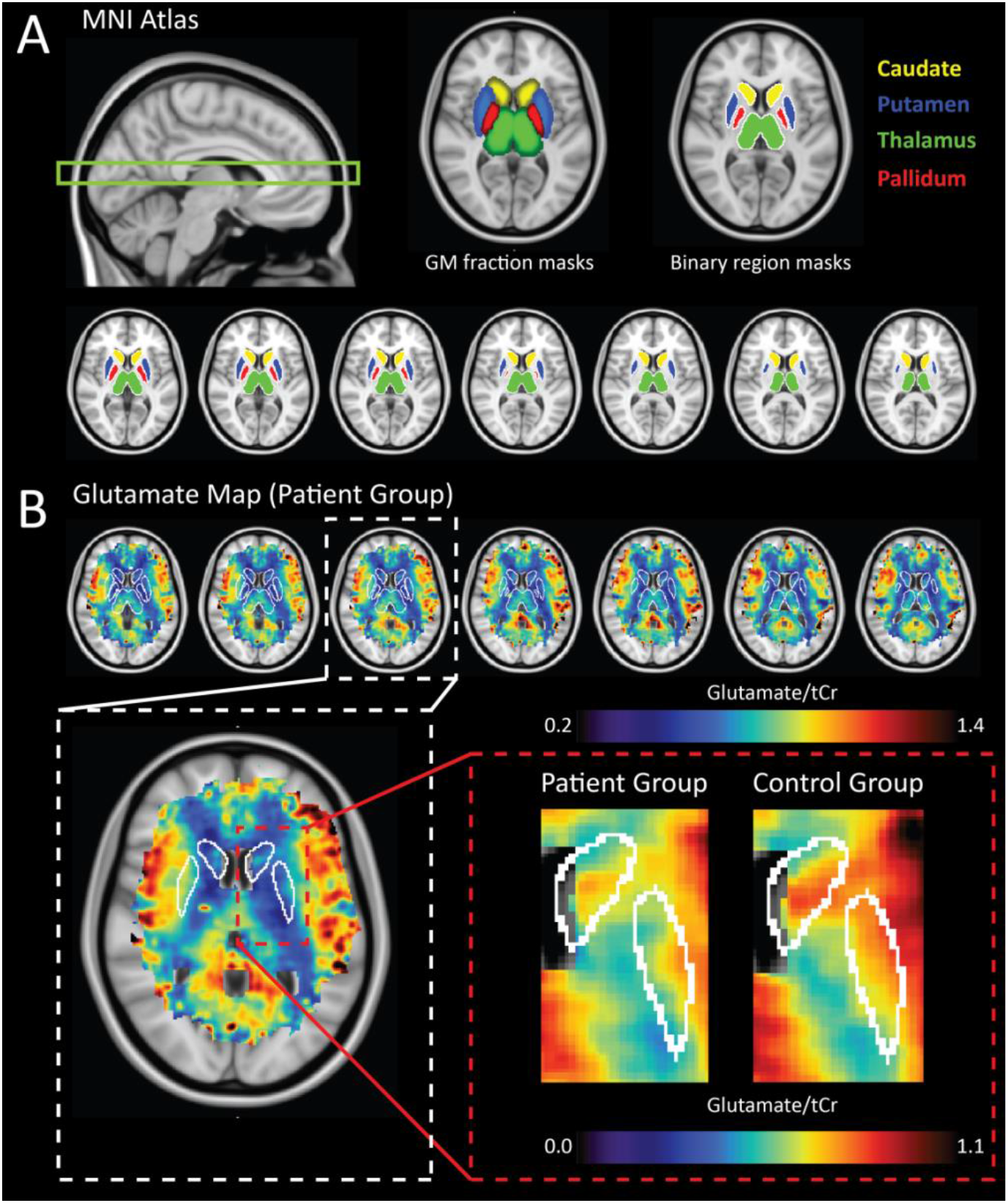
Deep GM regions were delineated based on tissue masks associated with the MNI standard atlas. GM tissue fraction maps (A-middle) were thresholded to create binary masks (A-right) of the Caudate (> 30%), Putamen (> 50%), Thalamus (> 60%) and Pallidum (> 10% GM). The target region for our acquisition is shown in the sagittal image (A-left). Transverse anatomical MNI images along with corresponding average Glu maps for the patient group are show (A-right); **1B-top**: selected transverse slices of the average Glu map from the patient group are shown along with delineated ROI; **1B-bottom**: zoomed images showing average data between patients (left) and controls (right) for the Caudate and Putamen, where significant reductions in Glu were observed. Figure scaling has been adjusted to highlight localized differences.

A 3D Turbo Field Echo (TFE) scan (TE/TR = 2.89/8ms, resolution = 1mm isotropic, FOV = 220×220×200 mm^3^, scan duration = 6min 51sec, flip angle = 6 degrees) and a shimmed dual echo GRE B_0_ map (delta TE/TR = 2.35/5.12 ms, FOV = 220×220×30 mm^3^, acquisition matrix = 176×176, resolution = 1.25 x 1.25 x 10mm^3^, slices = 3, scan duration, 1m 6s) and 2D multi-slice T1w Fast Field Echo (FFE) image (TE/TR = 4.22/200 ms, FOV = 220×220×30 mm^3^, acquisition matrix = 176×176, resolution = 1.25 x 1.25 x 10mm^3^, slices = 3, scan duration, 72 s) were acquired. The FFE and B_0_ map were used to perform over-discrete reconstruction with B_0_ correction,^29^ while the whole brain TFE anatomical image facilitated spatial normalization of MRSI data to the 1mm MNI atlas. Tissue segmentations (GM, WM and CSF) were used to perform partial volume and T1 correction of Glu maps. Full spectral reconstruction and post-processing as well as metabolite map and quality assurance map generation details as previously published.^24^ Region of interest masks of brain regions from which sufficient coverage was attained (i.e. the Caudate, Putamen, Thalamus and Pallidum) were generated using MNI tissue fraction maps (**Figure 1A**) using FSL (FMRIB).^30^ Resulting masks were binarized to create masks of the GM nuclei. The combined deep GM region was created by amalgamating these regions of interest.

### Statistical Analysis

To compare demographic characteristics, independent t-tests and chi-squared tests were performed. The average glutamate values in the combined deep GM region was compared between patients and controls using an independent samples *t*-test. A coverage threshold of 10% was used; below that, data was excluded from statistical analysis. Stratified analyses were subsequently performed on individual deep GM regions to assess whether any effects could be further localized to individual regions, again applying a 10% coverage threshold. Sensitivity analyses were performed using age-matched groups to exclude potential confounding effects of age. Additionally, to verify the generalizability of the pair-wise differences we had found at the 10% coverage threshold, we varied the threshold for sufficient signal coverage across subcortical brain areas from 5% to 35% with 5% increments and performed group-wise comparisons at each increment. For brain regions that differed between patients and controls, Spearman’s correlations between glutamate levels and potentially confounding variables were calculated, i.e. age and the defined daily dose (DDD) of antipsychotics, and t-tests were performed to assess differences between sexes. The relation between glutamate levels and overall cognition as well as individual cognitive domains was assessed using Spearman’s correlations corrected for age and using Bonferroni correction for the cognitive domains. These correlations were calculated across all participants (i.e. patients and controls) due to the simple size, although this that a relation between glutamate levels and cognition is the same within both groups. Statistical analyses were performed in SPSS (version 26.0.0.1). We checked normality using histogram inspection and *p*-values less than .05 were considered statistically significant. All results are reported as means ± standard deviation.

## RESULTS

### Participant characteristics

Demographics and clinical characteristics are displayed in **Table 1**. In total, 23 healthy control subjects were recruited from the general population (age 24±6 years, 9 females) and 16 individuals with a psychotic disorder (age 33±11 years, 4 females). The average age in patients was lower (*p*=0.008) but gender ratios were comparable (*p*=0.711). Eight patients were diagnosed with schizophrenia, six with psychosis not otherwise specified, and two with a schizoaffective disorder. As expected, overall BACS scores were lower in patients with a psychotic disorder compared to controls (t(37)=3.36, p=0.002). For the individual cognitive domains, scores were significantly lower for verbal memory (p=0.012) and attention/processing speed (p<0.001), but not for working memory, psychomotor speed, verbal fluency, and executive functioning (all p-values >0.05).

**Table 1.** Demographics and clinical parameters.

|  | Controls<br>(N=23) | Patients<br>(N=16) | Pairwise comparisons |  |
| --- | --- | --- | --- | --- |
|  |  |  | Test statistic | p-value |
| Age | 23.87 ($\pm 5.7$) | 33.13 ($\pm 11.7$) | $t(20)=-2.93$ | 0.008* |
| Gender (F/M) | 7/16 | 4/13 | $X^2(38)=0.14$ | 0.711 |
| Average cognition | 0.19 ($\pm 0.66$) | -0.76 ($\pm 0.81$) | $t(37)=-3.36$ | 0.002* |
| Verbal memory | 0.04 ($\pm 1.01$) | -1.50 ($\pm 2.06$) | $t(20)=2.76$ | 0.012* |
| Working memory | 0.06 ($\pm 1.04$) | -0.47 ($\pm 1.34$) | $t(37)=1.40$ | 0.196 |
| Psychomotor speed | 0.06 ($\pm 1.04$) | -0.56 ($\pm 1.12$) | $t(37)=1.779$ | 0.083 |
| Verbal fluency | 0.01 ($\pm 0.88$) | -0.37 ($\pm 0.72$) | $t(37)=1.432$ | 0.161 |
| Attention/processing speed | 0.02 ($\pm 1.00$) | -1.40 ($\pm 1.24$) | $t(37)=3.949$ | <0.001* |
| Executive functioning | 0.00 ($\pm 1.00$) | -0.26 ($\pm 0.79$) | $t(37)=0.882$ | 0.383 |
| Antipsychotics (Y/N) | - | 13/3 |  |  |
| Defined daily dose (n=6) | - | 0.76 ($\pm 0.41$) | | |
| Antidepressants | - | 4/12 |  |  |
| Mood stabilizers (Y/N) | - | 2/14 |  |  |
| Benzodiazepines (Y/N) | - | 5/11 |  |  |
| Diagnosis (SZ,PN,SA) | - | 8/6/2 |  |  |
| Age of onset psychosis | - | 23.67 ($\pm 10.1$) | | |
| Years since first psychosis | - | 10.71 ($\pm 11.5$) | | |

F=female, M=male, Y=yes, N=no, SZ=schizophrenia, PN=psychosis not otherwise specified, SA=schizoaffective disorder, <sup>a</sup>=one-sample t-test ( $h_0=1$ ).

### Glutamate levels in deep brain structures

Compared to healthy controls, overall glutamate levels in combined deep GM structures were significantly lower in patients with a psychotic disorder compared to healthy controls (0.67±0.07 vs. 0.75±0.08; *t*(32)=2.98, *p*=0.005, *d*=1.06) (**Figure 2, top**). Stratified analyses showed significantly lower glutamate levels in the caudate (0.70±0.11 vs. 0.62±0.10; *t*(31)=2.08, *p*=0.046, *d*=0.76) and putamen (0.75±0.08 vs. 0.66±0.11; *t*(31)=2.63, *p*=0.013, *d*=0.94), but not in the pallidum (0.56±0.14 vs. 0.54±0.12; *t*(14)=0.19, *p*=0.853, *d*=0.15) or thalamus (0.76±0.10 vs. 0.72±0.09; *t*(34)=1.29, *p*=0.206, *d*=0.42; **Figure 2, bottom**). Sensitivity analyses with different coverage signal thresholds (5-35%) showed that decreased deep GM glutamate levels were present across coverage signal thresholds, even though the statistical power due to decreasing sample size affected statistical significance to a certain degree (see Supplementary Figure S1). With increasing threshold, the variance of the two groups was reduced as more coverage would lead to a more reliable signal and reduce the possibility of outliers.

**Figure 2.**
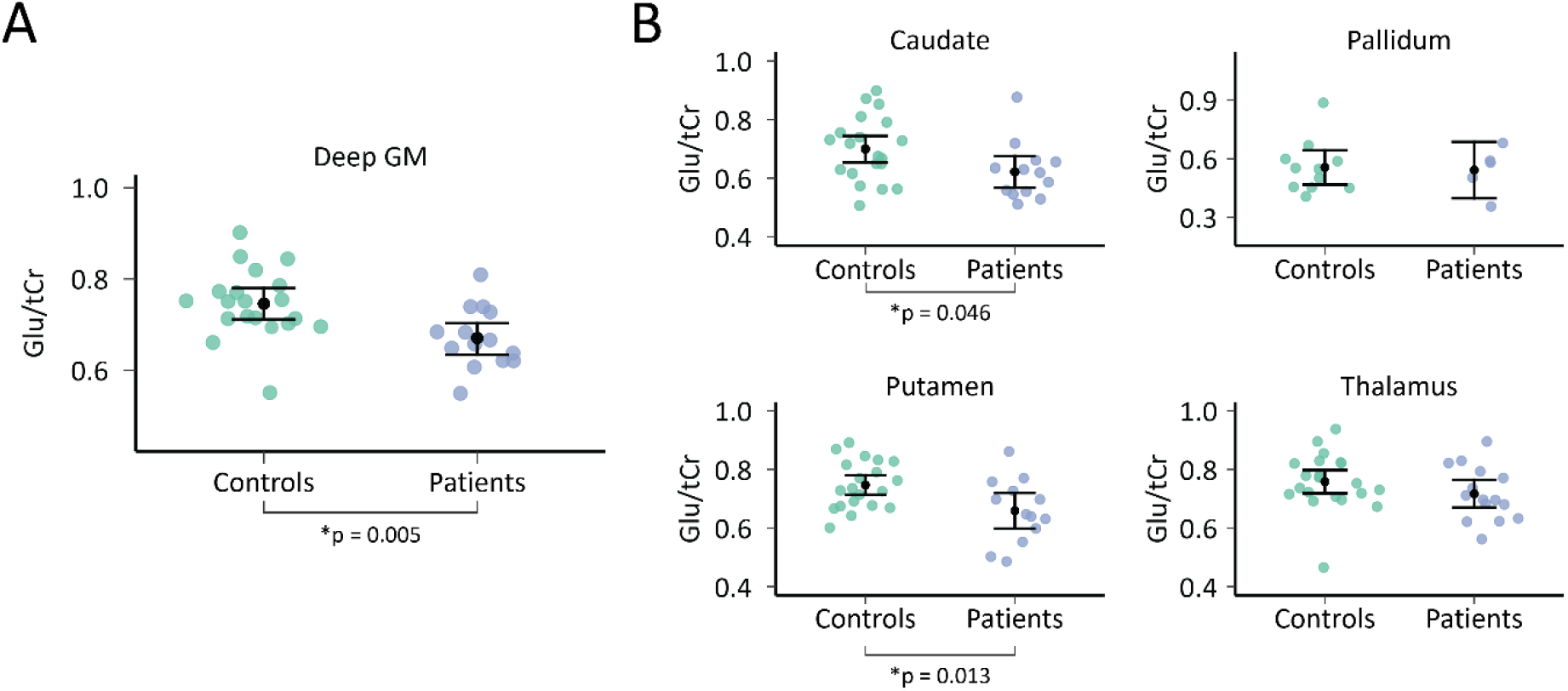
The Glu/tCr ratio found per subject in the control and patient groups. **Figure 2A:** the ratios were portrayed for the deep GM as a whole. **Figure 2B**: Glutamate levels per anatomically defined brain region with sufficient data to perform statistical analysis. Means and 95% confidence intervals per group are superimposed on the individual data points. Significant differences were found in whole deep GM, and the Caudate and Putamen (P<0.05). *Glu = Glutamate, tCr = total creatine, GM = gray matter*.

### Analysis of possible confounders

In both patients and controls, age was significantly related to glutamate levels in the combined deep GM (*r*(32)=-0.43, *p*=0.010) and caudate (*r*(31)=-0.36, *p*=0.041), but not in the putamen (*r*(30)=-0.25, *p*=0.169). Therefore, to ensure that the findings were not drive by age differences, patients (*n*=15) and controls (*n*=15) were matched by age (30 vs. 26 years, *t*(28)=-1.58, *p*=0.13). With comparable gender ratios (patients: 20% female, controls: 27% female, *X*^2^(28)=0.186, *p*=0.67), deep GM glutamate values remained significantly lower in patients (0.67±0.07) compared to controls (0.75±0.09; *t*(23)=2.383, *p*=0.026), including lower glutamate levels in the caudate (0.62±0.10 vs. 0.71±0.11; *t*(24)=2.143, *p*=0.042) and putamen (0.67±0.11 vs. 0.77±0.08; *t*(22)=2.588, *p*=0.017). No differences in glutamate values in the combined deep GM was found between men and women (*t*(32)=0.57, *p*=0.571), caudate (*t*(31)=-0.36, *p*=0.718), and putamen (*t*(31)=-0.14, *p*=0.892). Within patients, the DDD of antipsychotics was not significantly correlated to total deep GM glutamate levels (*r*(7)=-0.60, *p*=0.089), nor for the caudate (*r*(8)=-0.60, *p*=0.065), and putamen (*r*(6)=-0.293, *p*=0.482), although a negative trend was observed.

### Glutamate levels in relation to cognition

Overall **c**ognitive scores were significantly correlated to glutamate values across deep GM structures (*r*(31)=0.38, *p*=0.031; **Figure 3, left**). Stratified analyses did not show statistically significant correlations in the caudate (*r*(30)=0.32, *p*=0.071), putamen (*r*(30)=0.34, *p*=0.057), thalamus (*r*(33)=0.28, *p*=0.102), or pallidum (*r*(11)=-0.09, *p*=0.752). Subsequently, we analyzed individual cognitive domain scores in relation to combined deep GM glutamate levels. A significant relation was found between glutamate levels and psychomotor speed (*r*(31)=0.53, *p*_*corr*_=0.012; **Figure 3, right**), but not for verbal memory (*r*(31)=0.249, *p*_*corr*_=0.97), working memory (*r*(31)=-0.124, *p*_*corr*_=0.99), verbal fluency (*r*(31)=0.258, *p*_*corr*_=0.88), attention/processing speed (*r*(31)=0.310, *p*_*corr*_=0.48), and executive functioning (*r*(31)=0.237, *p*_*corr*_=0.92).

**Figure 3.**
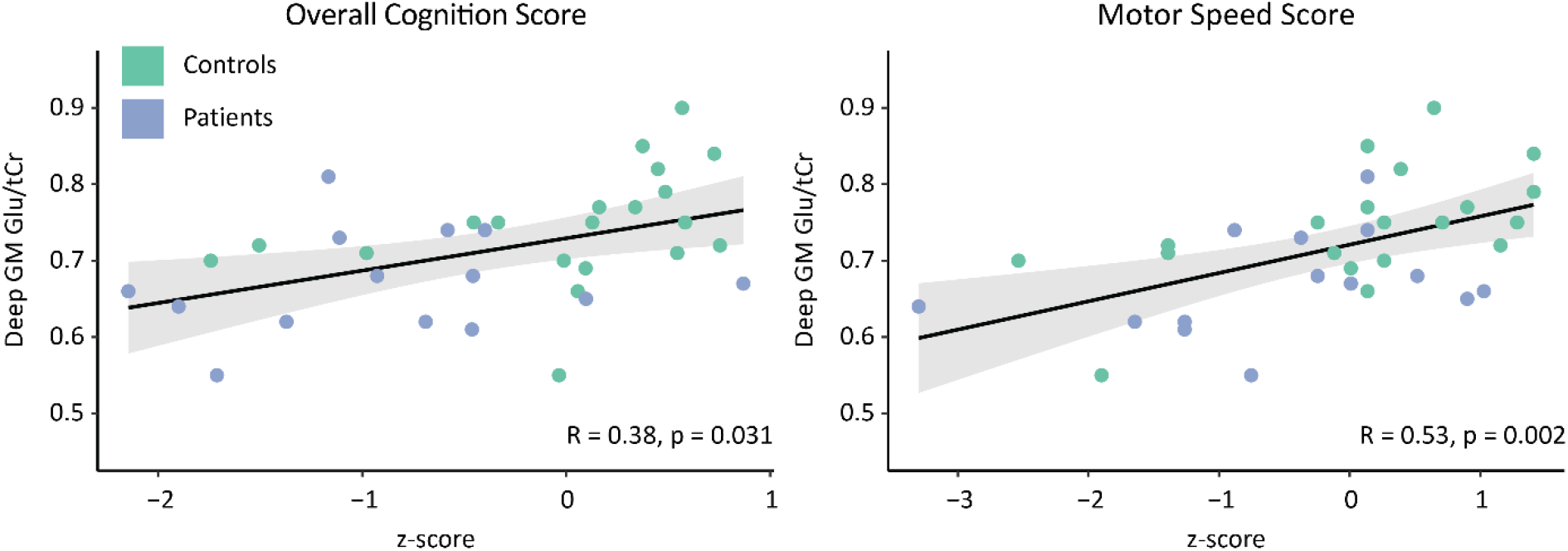
Deep gray matter glutamate levels positively correlate to the overall BACS score (left) and investigating individual cognitive domains showed that primarily psychomotor speed is related to glutamate levels in patients with a psychotic disorder and healthy controls (Spearman’s correlation, adjusted for age). The BACS cognition scores represent normalized values (i.e. z-scores). Glutamate values represents the glutamate to total creatine ratio. *Glu=glutamate, tCr=total creatine, GM=gray matter, BACS=brief assessment of cognition in schizophrenia*.

## DISCUSSION

Using advanced 2D MRSI techniques, this study found reduced glutamate levels in deep GM structures in patients with a psychotic disorder which was particularly pronounced in the caudate and putamen. We have found that deep GM glutamate positively related to cognitive performance, specifically to psychomotor functioning and not to verbal memory, working memory, verbal fluency, attention and processing speed, and executive function. Our approach included a high-resolution 7T acquisition with precise delineation of deep GM structures while mitigating sources of error through novel artefact control techniques and correction for tissue-specific GM and WM glutamate contributions. This is the first study to report the quantification of glutamate across anatomically delineated deep GM regions simultaneously in patients with a psychotic disorder; in contrast to single-voxel spectroscopic measurements.

In support of our findings, studies employing higher magnetic field strengths have generally reported reduced glutamate levels in schizophrenia patients,^17-21, 31^ even though these studies generally focused on cortical areas (anterior cingulate cortex or occipital cortex) rather than deep brain structures. Several cellular mechanisms are possible through which glutamate levels can be altered in psychotic disorders. Blocking the NMDA receptor through ketamine administration in healthy humans has shown to increase glutamate^32^ and glutaminase,^33^ the key enzyme to convert glutamine to glutamate. Changes in the NMDA receptor or glutaminase activity might cause a shifting glutamate/glutamine balance.^10^ Our finding of overall reduced glutamate levels contrasts with a recent meta-analysis finding of elevated Glx signal in patients with schizophrenia,^16^ but very few studies have quantified bilateral glutamate levels across deep brain structures. Moreover, it is important to note that MRSI-studies performed at lower magnetic field strength may be hindered by lower signal-to-noise ratios, greater partial volume effects due to large voxel size and more ambiguities in signal quantitation due to overlapping glutamate and glutamine resonances.

Cognitive deficits have a pronounced effect on the quality of life and functional outcome of patients with a psychotic disorder.^1, 2^ We found that glutamate levels are involved in cognitive function related to psychomotor outcomes. In general, these results complement a previous 4T-MRSI study,^7^ as that study found that cortical glutamate levels positively related to a range of cognitive domains in schizophrenia patients. Although overall cognition was impaired in our patient sample, psychomotor speed was not. Nevertheless, psychomotor speed problems are a well-known phenomenon in patients that have experienced a psychosis.^34^ Deep GM regions, including striatal structures (e.g. putamen and caudate), are involved in motor actions.^35^ Moreover, previous research has shown that the putamen was involved in psychomotor speed,^36^ supporting the finding that altered glutamate levels in these regions could relate to altered psychomotor functioning.

Notwithstanding important methodological improvements, our study has several limitations. First, our sample size is limited and consisted of patients that are not antipsychotic-naÏve with dissimilar disease durations. This is important as antipsychotic drugs may influence glutamate levels,^37^ and antipsychotics have effects on psychomotor speed and may even exacerbate cognitive problems through dysregulation of N-methyl-D-aspartate (NMDA) receptor activity.^38^ Similarly, a better understanding is needed whether glutamate levels in deep gray matter structures are decreased already prior to treatment and whether the same regions display glutamate level alterations before and after treatment. Second, in order to remove extracranial lipid signal contamination, we explicitly focused on deep brain structures to improve the accuracy of metabolite quantification, but this comes at the cost of cortical glutamate signals for which analyses were not possible due to the use of extracranial signal suppression. Finally, creatine was used as internal reference, in line with current literature,^39^ to divide out deviations related to coil loadings and inhomogeneous B1 transmit/receive fields. Absolute quantitation might further improve the accuracy of the reported metabolite levels in future studies.

As technological innovations expand the applicability of MRSI as a method to evaluate neuronal metabolism, future studies will benefit from emerging techniques for whole-brain metabolic imaging. This necessity is underscored by our observation that glutamate levels in deep GM structures primarily related to psychomotor speed functioning, while cortical glutamate, as measured by Bustillo et al, related to broader cognitive domains ^7^. A whole-brain approach would make it possible to localize specific metabolic consequences and relate them to associated known functional domains simultaneously to generate cognitive profiles. By further accelerating spectroscopic imaging techniques at high resolutions, functional magnetic resonance spectroscopy can also provide insights into the dynamic aspects of neuronal metabolism that may also be altered due to psychiatric disease.^40^

## Conclusion

Our results highlight the value of applying high resolution MRSI to understand cognitive deficits related to psychotic disorders by reliably quantifying glutamate levels across anatomically defined subcortical brain structures. Moreover, our study shows that novel methodological methods to measure detailed levels of *in vivo* glutamate across deep brain structures are important tools to understand the mechanisms of psychotic disorders.

## Data Availability

Raw data is available from the authors upon reasonably justified request.

## Acknowledgements

This research was supported by a Brain and Behavior Research Foundation NARSAD Young Investigator Award (Christiaan H. Vinkers, 24074).

## Author Contributions

TAAB, AAB, and CHV made substantial contributions to the conception or design of the work. All authors were involved in drafting the work or revising it critically for important intellectual content and gave final approval of the version to be published. Finally, TAAB, AAB, and CHV are in agreement to be accountable for all aspects of the work in ensuring that questions related to the accuracy or integrity of any part of the work are appropriately investigated and resolved.

