## supplemental for "Glutamate levels across deep brain structures in patients with a psychotic disorder and its relation with cognitive functioning"

**SUPPLEMENTARY INFORMATION**

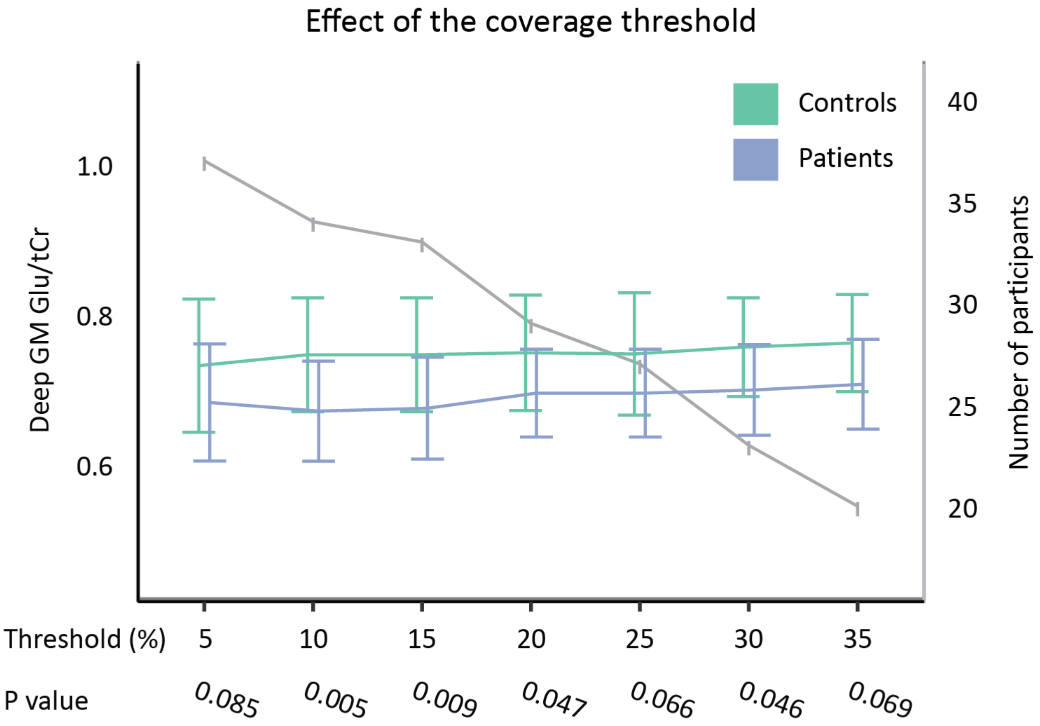


**Supplementary Figure S1.** The effect of the coverage (i.e. the percentage of deep brain structure covered by the MRS-signal) threshold on glutamate level differences between patients with a psychotic disorder and healthy controls. The x-axis represents the coverage threshold. The blue and green lines represent the Glu/tCr values in patients (blue) and controls (green; left y-axis), at the same time the grey line shows the number of patients remaining after thresholding (right y-axis). The means were calculated for every 5% increment and plotted with slight offsets around the threshold for better visualization, the error bars represent the standard deviations. Glu = Glutamate, tCr = Total Creatine, GM = gray matter.
